# Influenza Outbreak Prevention and Management in U.S. Carceral Settings: A Scoping Review

**DOI:** 10.1101/2022.02.06.22270517

**Authors:** Max Jordan Nguemeni Tiako, Julia Zubiago, Yvane Ngassa, Rubeen Guardado, Nicolas Munoz, Alysse G Wurcel

## Abstract

**Background:** Detention settings’ preparedness against respiratory virus outbreaks is essential, with implications for preventing illness and deaths from future pandemics. We sought to identify influenza outbreak prevention and management evidence in U.S. detention settings.

**Methods:** We conducted a scoping review, first searching *PubMed, OVID, Google Scholar, Medline* databases and the reference lists of identified manuscripts published on outbreak prevention and management of influenza in detention settings in English. Search terms included *prison, jail, vaccine, influenza, outbreak, management, prevention, carceral*.

**Results:** Twenty-five studies met the search criteria, ultimately narrowed down to seven studies. Four studies focused on prevention and three on management. The studies on prevention identified restructuring housing, vaccinations, and widespread screening to prevent outbreaks and highlighted the importance of collaboration between prison staff and public health departments. The management studies emphasized hygiene, isolation of sick individuals, and vaccination of unexposed patients. Staff expressed the concern that the public may view prisoners as low priority based on prior experiences with influenza vaccine shortages, with a spillover effect in obtaining vaccines and medications for staff. No studies mentioned decarceration as a prevention and mitigation measure.

**Conclusion:** There is limited data on influenza outbreak prevention and management in detention settings. The approaches described are partially in line with public health recommendations but fall short due to lack of, and delays in resource allocation. There is an urgent need for researchers and public health officials to examine and report influenza outbreak prevention and mitigation strategies in detention settings to develop scalable interventions and a national standard for all detention settings.

## Introduction

Prisons, jails, and immigration detention centers are epicenters of the transmission of SARS-CoV-2 and the continued spread of the COVID-19 pandemic.^1–3^ Nearly 400,000 people in state and federal carceral settings have tested positive with COVID-19, and over 2,000 people who are incarcerated have died.^2^ Several studies have documented higher case rates and test positivity among detainees in immigration detention centers,^4^ prisons, and jails.^5^ Similarly, the death rate in prisons and jails has been higher than the U.S. population.^3,6^ Detained individuals carry a greater burden of chronic diseases that put them at an increased risk of severe morbidity and mortality should they contract COVID-19.^7,8^ The spread of COVID-19 in carceral settings is linked to community spread as evidenced by a study from Chicago that found jail cycling (being incarcerated and then being released to the community) was a significant predictor of SARS-CoV-2 infection.^9^

The incarcerated population in the U.S. includes disproportionately Black, Hispanic, and Indigenous people. Black, Hispanic and Indigenous communities have been significantly affected by the COVID-19 pandemic in terms of infection rates and mortality due to occupational and residential risk, and prevalence of chronic illnesses associated with increased COVID-19 mortality risk.^10,11^ The disproportionate impact of mass incarceration and the COVID-19 pandemic on communities of color in the U.S. highlights the need for public health efforts to focus on addressing the risk of infectious spread among incarcerated people and individuals work in carceral settings.

Little exists on U.S. jails, prisons, and detention centers’ preparedness and ability to prevent and mitigate outbreaks of respiratory viruses such as SARS-COV-2. While the SARS-COV-2 virus is relatively new, this information may be inferred from evidence on carceral settings’ handling of influenza outbreaks, given the frequency and seasonality of influenza infection rates. We, therefore, sought to conduct a scoping review of the published literature on the prevention and management of influenza outbreaks in U.S. prisons, jails, and detention centers to inform future approaches to infection control in carceral settings.

## Methods

### Search strategy

We used a scoping approach to identify papers about preventing and managing influenza outbreaks in detention centers. Two authors (MJNT, JZ) searched *PubMed, OVID, Google Scholar, Medline* and the reference lists of identified papers and completed the triaging process. Our search terms included *prison, jail, vaccine, influenza, outbreak, management, prevention, carceral*. We restricted our search to English-language studies. We included studies that reported data on measures of prevention and management of influenza outbreaks and excluded secondary surveys of infectious diseases data, perspectives, and opinion pieces. We restricted ourselves to studies of U.S. carceral settings. Two authors (MJNT & JZ) separately reviewed the titles and abstracts for relevance based on pre-established eligibility criteria (Figure 1).

**Figure 1:**
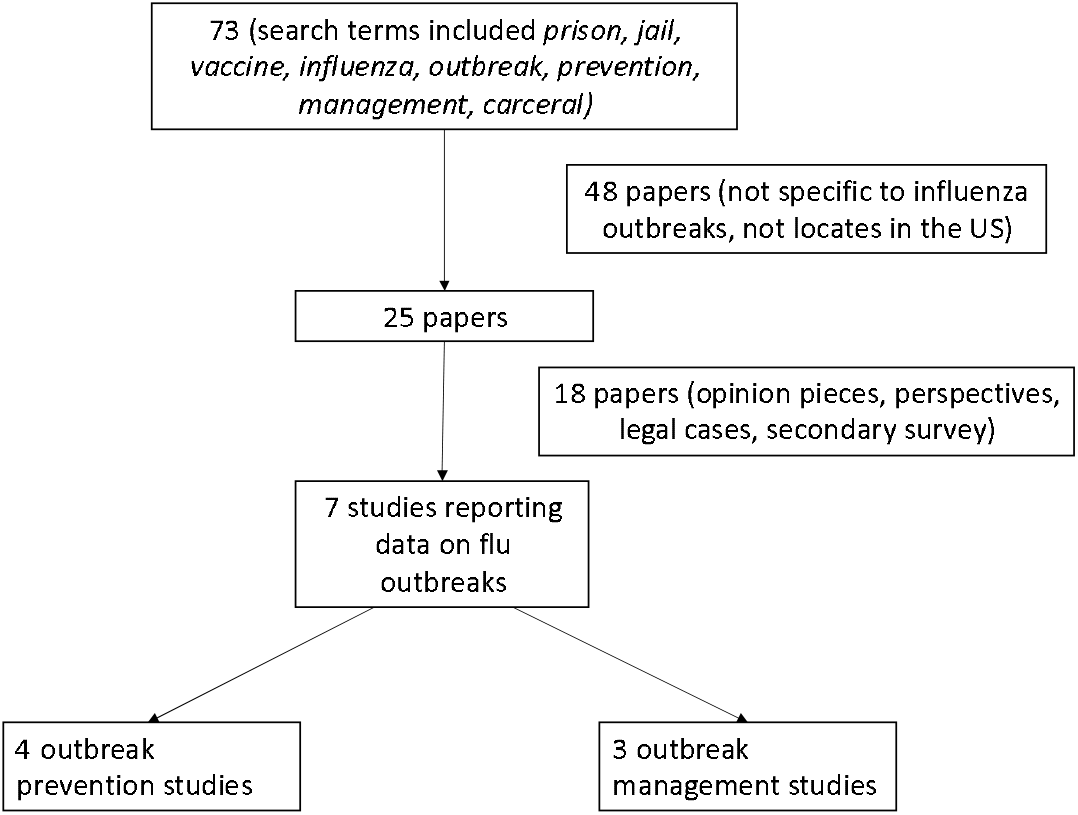
Flow chart of the literature search and study selection process in a review of the literature on influenza outbreak prevention and management in U.S. prisons and Jails

### Data extraction

Three authors independently reviewed and extracted the relevant data (NM, JZ, MJNT), including 1) the years during which the data were collected and the study design, 2) the type of data collected, the key topic covered (prevention vs. outbreak management) 3) the location of the study, 4) characteristics of the study (population size, type of carceral setting) and 5) main outcomes and future recommendations from the study.

## Results

There were a total of 25 studies that met the original search criteria, further narrowed down to seven studies. Of the seven studies that met review criteria, we categorized as four outbreak prevention, and three as outbreak management (Table 1).

**Table 1:** Review of Influenza Outbreak Prevention

|  | Article Title | Authors | Year | Journal | Study Location | Study Type |
| --- | --- | --- | --- | --- | --- | --- |
| Prevention | Distribution of A(H1N1)pdm09 Influenza Vaccine: Need for Greater Consideration of Smaller Jails | Lee et al | 2014 | Journal of Correctional Health Care | USA | Cross-Sectional Study with mixed-methods analysis |
|  | How public health and prisons can partner for pandemic influenza preparedness: a report from Georgia | Spaulding et al | 2009 | Journal of Correctional Health Care | Georgia, USA | Descriptive Study |
|  | Pandemic influenza and jail facilities and populations | Maruschak et al | 2011 | American Journal of Public Health | USA | Descriptive Study |
|  | Influenza Vaccination in Massachusetts Jails: A Mixed-Methods Analysis | Khorasani et al | 2021 | Public Health Reports | Massachusetts, USA | Survey, mixed-method study |
| Management | Influenza outbreaks in two correctional facilities -- Maine, March 2011 | Robinson et al | 2012 | CDC Morbidity and Mortality weekly report | Maine, USA | Epidemiological Investigation Report/Case Study |
|  | Pandemic influenza as 21st-century urban public health crisis | Bell et al | 2009 | Emerging Infectious Diseases | New York, USA \| Mexico City, Mexico | Descriptive Study |
|  | Responding to an H1N1 outbreak in an urban jail setting: The NYC experience | Erik Berliner | 2009 | Corrections Today | New York, USA | Descriptive Study |

### Outbreak Prevention

Spaulding^12^ reported that participating in interactive scenarios where medical staff at Georgia prisons and jails practiced strategies to manage influenza outbreaks led to an improvement of staff knowledge of preventative strategies (69% of medical staff correctly answered questions about hygiene, PPE, and influenza transmission post-exercise, compared to 42% pre-exercise). The medical staff identified several challenges to adequate pandemic influenza preparedness in prisons. Challenges included structural limitations on housing, preventing person-to-person spread, limited healthcare surge capacity in prisons (limited personnel, constrained medication availability and on-site distribution, reliance on outside hospitals for critical care, lack of redundancy in the system, and low morgue resources). Staff expressed the concern that the public may view prisoners as low priority based on prior experiences with influenza vaccine shortages, with a spillover effect in obtaining vaccines and medications for staff.

Maruschak^13^ conducted a descriptive study looking at data on jails from the U.S. Bureau of Justice Statistics (BJS) to analyze the unique challenges of influenza prevention in the jail setting. Over a third (1,100) of jails in the U.S. hold fewer than 50 inmates on an average day, while the 170 largest jails hold about 52% of the U.S. jail population, averaging more than 1,000 people in jail per day. Smaller jails had higher turnover rates than larger jails (100% vs. 54%/week) and operated at lower capacity than larger jails (67% vs. 100%). Fewer than 10% of the jails in the U.S. are accredited by organizations that have health standards for infection control. The research team recommended re-organizing dorm or cell spaces to prevent influenza transmission among the people churning between the jail setting and outside. The research team recommended re-organizing dorm or cell spaces to limit transmission and designating quarantine areas. The authors also recommended that planners develop a strategy to screen inmates and staff for influenza accounts for potential resource shortages during a pandemic.

To model the predictors of administering pandemic influenza vaccine, Lee^14^ conducted a cross-sectional survey with qualitative and quantitative sections at over 1000 correctional facilities (31 federal prisons, 341 non-federal prisons, and 814 jails) during the H1N1 pandemic. Small jails were the least likely to report having received influenza vaccine (44%) and having an influenza plan (41%) compared to larger jails. There was a 32% greater likelihood of receiving the influenza vaccine for every 100 inmates increase in the daily population. Jails that reported a pandemic plan had a 69% greater likelihood of receiving the influenza vaccine. Respondents identified that their biggest concerns were the late arrival of vaccines and limited quantities of vaccines. The lessons reported from this study included the need to secure vaccines and vaccinate early, develop precautions, educate inmates and staff about influenza, and communicate with the public and public health authorities.

Khorasani et al.^15^ conducted a mixed-methods survey study in 14 Massachusetts jails’ health services administrators (HSAs) about institutional influenza vaccine distribution practices and policies in jails. They also calculated influenza vaccination rates for each jail facility using influenza vaccine orders data from the Massachusetts Department of Public Health (DPH). Eleven (79%) health service administrators in M.A. responded to the survey; all respondents reported the influenza vaccine was available to people pretrial, sentenced, and pre-released and reported high satisfaction with the vaccine distribution at their jails. However, only 72% of HSAs reported awareness of influenza vaccine policies at their jail facility. There were no significant differences among jails in influenza vaccine policies and practices. The authors found low influenza vaccination rates in Massachusetts jails (<10%) and emphasized the need to create a database that reflects the true vaccination rates in jails.

### Outbreak Management

Berliner^16^ described the Department of Corrections (DOC) and Department of Health and Mental Hygiene (DOH) response to a confirmed case of H1N1. The DOC and DOH focused on controlling the spread through preventing close-quarters socializing between newly admitted inmates and others, separating the known exposures from unexposed, and using teleconferencing for court visits. After the first case of H1N1 was confirmed, the housing unit was placed on quarantine, and anyone with symptoms was transferred to the DOC’s infectious disease unit, with negative pressure isolation cells. The staff were given personal protection equipment and taught to screen for symptoms among all inmates and new inmates at intake or transfer. Eleven additional inmates were identified as possible cases during the first week and transferred to the DOC infectious disease units. Overall, 63 housing areas were placed on medical restriction (1,000 inmates at peak). All pregnant female inmates were moved to one housing area and placed under precautionary restrictions.

Bell^17^ described the city-wide efforts in New York City to control the spread of H1N1 pandemic influenza. NYC used a novel surveillance system to monitor emergency department visits, looking for increased influenza symptoms as the chief complaint. These data were transmitted to the DOH to monitor the community’s spread. Once cases were confirmed, an organization called *Ready New York* spearheaded multilingual public outreach to minority communities, messaging around hand hygiene, avoidance of possibly sick people, and social distancing.

Robinson^18^ wrote a CDC Morbidity and Mortality Weekly Report describing an outbreak in two prisons in Maine. One was a medium- to maximum-security prison (A), and the other was a minimum-security prison (B). An inmate was admitted to the hospital with influenza. When another tested positive, the Maine CDC sent public health nurses to assess influenza-like illness and give treatments to patients. Facility A screened 802 people, gave treatment to the 2.1% who reported symptoms, and vaccinated over 300 people. 184 staff at facility A were screened, 18% received treatment for symptoms, and 37% were vaccinated. Facility B screened 193 people (2.1% received treatment for symptoms) and vaccinated 46% of their population. Among staff, 51 were screened (17.6% of staff received treatment for symptoms), and 25% were vaccinated. The authors highlight the collaboration between public health and corrections officials to identify and mitigate disease spread.

## Discussion

Our analysis has three main findings. First, the publications presenting data on influenza prevention identified primarily restructuring housing, widespread screening, and court teleconferencing as ways to contain epidemics and highlighted that collaboration between staff at carceral sites and public health departments is crucial. Second, the available literature in U.S. carceral systems focused on institutional or city-wide experiences, with largely reactive rather than proactive planning, with a low emphasis on prevention. Lastly, there was a lack of evidence on this important topic.

That so little research focuses on health in carceral settings may be multifactorial. First, studies involving incarcerated populations require additional IRB scrutiny,^19,20,^ which impacts the scope of these studies and likely researchers’ interest in studying incarcerated populations, given the additional administrative burden. However, research, such as on preventing and mitigating influenza outbreaks in prison, aligns with directly improving the health or well-being of individual prisoners. Still, we found no data on influenza vaccination rates in U.S. carceral facilities. The only national organization for credentialing healthcare in jails—National Commission on Correctional Health Care (NCCHC) standards “require that incarcerated people be provided with clinical preventive services, including flu shots and other immunizations administered as clinically indicated.”^21^ In the context of COVID-19, the Centers for Diseases Control and Prevention (CDC) encourage prioritizing incarcerated populations for COVID-19 vaccination, in line with historical recommendations that incarceration is a period when detention centers should offer vaccinations.^22^ While this guidance applies to prisons; there are no federal, state, or local guidelines for vaccine delivery in jails. Vaccination rates in jails are suboptimal. Indeed, a recent study of people in correctional and detention settings in 4 states showed that only 45% of people were willing the get vaccinated against covid-19.^23^ However, there is little research on how to improve the process of vaccine delivery and uptake in such settings, highlighting the need for greater transparency in vaccine policies and protocols in carceral settings. Influenza vaccination uptake data, including attitudes towards vaccines, may be particularly helpful in the rollout of the COVID-19 vaccine. Given the long history of government-sanctioned abuses on minority populations both in research and treatment settings,^24^ the long history of and lingering segregated healthcare provision and healthcare discrimination,^25^ vaccine hesitance may be particularly high among minorities in state detention. Evidence shows that mistrust in government influences Black people’s lower confidence in the influenza vaccine.^26^ This mistrust is likely exacerbated among individuals in government custody, especially in the short term (given that they have even fewer incentives to partake in jail-level public health efforts), as recent evidence shows negative experiences with the criminal justice system decrease trust in medical institutions.^27^

Reported mitigation measures such as restructuring housing in detention centers, isolation, screening are in line with prior evidence and recommendations of influenza outbreak mitigation.^28,29^ They are also relevant in the context of the COVID-19 pandemic, as a statewide study showed that the majority of outbreaks and deaths occured within a handful of prisons, attributable to overcrowding.^30^ Similarly, widespread screening as reported is an important strategy to detect ad mitigate outbreaks. A CDC study of 16 prisons and jails showed that Mass testing resulted in a median 12.1-fold increase in the number of known infections among incarcerated or detained persons, highlighting the limitations of symptom-based testing for respiratory viruses.^5^

None of the papers identified decarceration as a method to decrease influenza transmission. Most Americans are much more supportive of harm reduction measures like improved sanitation than of releasing people from prisons and detention centers. Only one-third to one-half of Americans believe that response to COVID-19 in prisons and immigrant detention centers should be a high priority, in line with correctional officers’ perception that the health of people who are incarcerated is a low priority to the public.^31^

The spread of SARS-COV-2 in U.S. detention centers highlights the need for mitigation measures such as decarceration,^32–34^ given the role crowding has on infectious spread in carceral^30^ and non-carceral settings.^35^ Decarceration has been a successful strategy used by several jails and prisons to decrease the spread of COVID-19. States and criminal justice officials have taken actions to reduce overcrowding in correctional facilities, such as the release of inmates, reduction of admissions, and sentence commutations.^36^ Nonetheless, living conditions in prisons and jails (such as crowding, suboptimal access to soap and water, and barriers to accessing timely healthcare) put people who are incarcerated at risk of infectious diseases.^1,37,38^ The health of people who are incarcerated directly impacts people who are not incarcerated. The churn between correctional facilities and communities of origin has implications for urban areas and the rural regions with limited healthcare resources but high concentrations of the nation’s correctional facilities, with implications for spreading around surrounding communities.^39,40^

Taken together, correctional officers’ perception that those who are incarcerated are a low priority to the public, the lack of data on influenza vaccinations in carceral settings, and the general scarcity of evidence on influenza outbreak prevention and management speak to the role structural racism plays in shaping both the experience of people who are incarcerated and knowledge production in the U.S. Indeed, The U.S. incarcerates disproportionately Black and Hispanic people,^41^ and the majority of immigrants in detention centers are Black and Hispanic and subject to longer detention times.^42,43^ Furthermore, scientists who study community health and health equity are significantly less likely to be awarded federal funding for their research.^44^

The main limitation of our study is the low number of papers we found that fit inclusion criteria. The strength of our work is that it is the first to appraise the available literature critically and can be used to encourage researchers to partner with carceral institutions and fill gaps in knowledge to inform the development of successful influenza vaccination and mitigation policies.

## Conclusion

The available literature reports on institutional or city-wide strategies for influenza prevention, with successful interventions built with collaborations between correctional officers and local public health officials. However, there remains a dearth of reporting and research on this subject. Outbreak prevention and mitigation approaches described in the literature partially align with existing public health recommendations but fall short due to lack of and delays in resource allocation. There is an urgent need for researchers and public health officials to examine and report on influenza outbreak prevention and mitigation strategies in detention settings, including influenza vaccination rates, to develop scalable interventions and a national standard for all detention settings.

## Data Availability

All data produced in the present study are available upon reasonable request to the authors

